# Characterizing and Predicting End-of-Life Patient Trajectories Using Routine Clinical Data

**DOI:** 10.1101/2025.08.11.25333434

**Authors:** Jonah Bosserhoff, Julius Keyl, Tim Lenfers, Dagmar Führer-Sakel, Marc Wichert, Frederick Klauschen, Martin Schuler, Sylvia Hartmann, Philipp Keyl, Jens Kleesiek

## Abstract

Understanding the biological processes that precede death is critical for making informed clinical decisions and facilitating care transitions. Here, we analyzed routine clinical data of 292,576 patients from two large hospital cohorts in Germany and the United States to identify temporal patterns at the end of life.

Integrating comprehensive laboratory values, vital signs, and ICD codes, we identified consistent, multisystem trajectories of terminal decline that were conserved across age, sex, and diagnostic subgroups. Changes largely occurred in an orchestrated pattern, beginning with electrolyte imbalances, followed by hepatic and renal dysfunction, progressive vital sign deterioration, and culminating in coagulation failure. Based on internal data from over 80,000 deceased and non-deceased patients, we developed a diagnosis-agnostic machine learning model to predict 90-day mortality risk using 27 routine clinical parameters and 15 disease groups. The prediction model had a high accuracy on internal data (AUC: 0.86) and generalized well to an external cohort of 211,527 hospitalized patients (AUC: 0.79).

Our results provide a data-driven foundation for understanding end-of-life pathophysiology and demonstrate the potential of routine hospital data to inform individualized care planning.

## Introduction

Although death is a fundamental aspect of medicine, its clinical characteristics remain poorly understood.^1,2^ While the medical field has traditionally prioritized the prevention and treatment of disease, the pathophysiological processes that precede death have received comparatively little systematic attention.^2,3^ However, a deeper understanding of end-of-life pathophysiology could be essential for distinguishing treatable conditions from irreversible decline and thereby help to guide the shift from curative to supportive care. This could support ethical clinical decision-making by complementing patient and clinician perspectives with objective biological evidence. Today, most patients in high-income countries die in hospitals, often following prolonged disease-oriented treatment and intensive monitoring.^4–6^ Advances in hospital infrastructure and the widespread adoption of electronic health records have led to the routine collection of large quantities of clinical data, including diagnostic codes, laboratory results, vital signs, and treatment histories.^7^ These rich, longitudinal datasets enable the systematic analysis of individual patient trajectories and population-level trends.^8^ They allow for fine-grained, disease-specific investigations, as well as the identification of shared pathophysiological patterns preceding death. This opens the door to a more nuanced understanding of terminal decline and the development of data-driven, tailored approaches to end-of-life care. In this study, we have analyzed two large real-world datasets integrating multimodal data, including laboratory values, ICD codes, and vital parameters, with a focus on their temporal evolution during the last year of life. To gain a comprehensive understanding of end-of-life pathophysiology, we included all documented deceased patients at each site, allowing for an inclusive, population-level view of terminal decline across diverse clinical settings. Our primary cohort consists of 42,508 deceased patients treated at a major German university hospital. To assess the generalizability of our findings, we compared results to an independent cohort of 37,525 patients derived from the publicly available MIMIC-IV database.^9–11^ In addition, we developed a machine learning model to predict patient outcomes at individual time points solely based on routine diagnostic parameters. Importantly, being disease-agnostic, our model is designed to capture generalizable features of terminal decline across diverse patient populations.

Together, this work demonstrates the potential of large-scale real-world data to uncover markers associated with end-of-life pathophysiology. It provides the foundation for data-driven decision support tools that can inform individualized care in advanced disease and critical care settings.

## Results

### Cohort description

We retrospectively evaluated clinical data from 42,508 deceased patients from University Hospital Essen and 37,525 deceased patients from the publicly available MIMIC-IV database (**Fig.1 + Suppl. Fig. 1**). The mean age of the Essen cohort was 66.3 years, with 41.2% female patients, while the MIMIC cohort had a slightly older mean age of 69.4 years and included 47.4% female patients. Data density differed between the cohorts, with an average of 878.13 features per patient in Essen, compared to 620.11 features in MIMIC.

**Figure 1:**
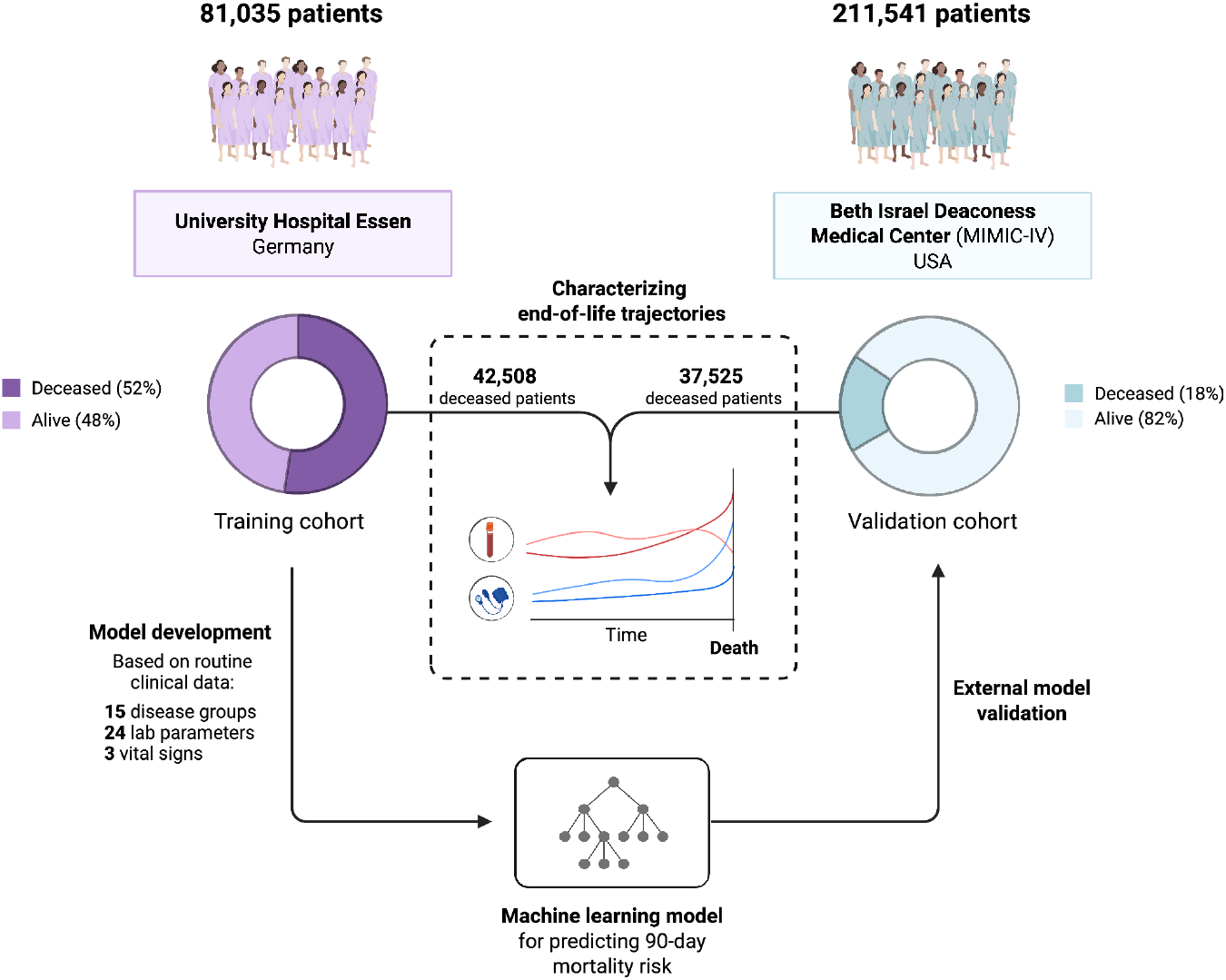
Overview of patient cohorts and study workflow

The most common ICD-coded diagnoses in the Essen cohort were neoplasms (n = 29,880), cardiovascular diseases (n = 18,923), and electrolyte imbalances (n = 17,715, **Table 1**). In contrast, the MIMIC cohort was dominated by hypertensive disorders (n = 27,426), cardiovascular diseases (n = 25,917), and electrolyte imbalances (n = 22,898). While the relative prevalence of specific conditions varied between cohorts, all major disease groups were comparably represented in both datasets.

**Table 1:** Characteristics of the deceased patient cohorts.

|  | Essen | MIMIC-IV |
| --- | --- | --- |
| Number of patients | 42,508 | 37,525 |
| Age (mean) | 66.3 | 69.4 |
| Age range | 18 - 106 | 18 - 106 |
| Sex (% female) | 41.2 | 47.5 |
| Features per patient (mean) | 878.13 | 620.11 |
| **ICD CODES** |  |  |
| Neoplasms | 70.3% (n=29,880) | 32.8% (n=12,322) |
| Cardiovascular Diseases | 44.5% (n=18,923) | 69.1% (n=25,917) |
| Electrolyte Imbalances | 41.7% (n=17,715) | 61.0% (n=22,898) |
| Hypertensive Disorders | 41.2% (n=17,526) | 73.1% (n=27,426) |
| Hematologic Disorders | 39.4% (n=16,750) | 58.0% (n=21,767) |
| Kidney Diseases | 33.0% (n=14,016) | 58.2% (n=21,857) |
| Pleural Effusion and Ascites | 25.4% (n=10,777) | 21.6% (n=8,109) |
| Liver Diseases | 20.4% (n=8,671) | 19.2% (n=7,199) |
| Diabetes Mellitus | 19.0% (n=8,085) | 33.1% (n=12,436) |
| Pneumonia | 19.0% (n=8,070) | 28.5% (n=10,698) |
| Sepsis | 18.1% (n=7,707) | 25.2% (n=9,452) |
| Obesity | 16.4% (n=6,958) | 54.1% (n=20,295) |
| Shock | 16.4% (n=6,984) | 11.2% (n=4,207) |
| Chronic Respiratory Diseases | 13.7% (n=5,819) | 28.1% (n=10,552) |
| Lymphomas | 10.7% (n=4,555) | 6.4% (n=2,398) |

### Progressive deterioration in laboratory and vital sign parameters precedes death

A wide range of clinical parameters displayed consistent and progressive changes during the final year of life. Markers such as lactate, lactate dehydrogenase (LDH), urea nitrogen, and white blood cell count steadily increased before death, while albumin, hemoglobin, and blood pressure decreased. These trends were highly consistent across the Essen and MIMIC datasets.

Notably, not all parameters demonstrated meaningful temporal changes. For example, body temperature remained largely unchanged throughout the observation period and did not show a significant trend prior to death on a cohort level. While some parameters, such as creatinine and lactate, showed a distribution shift between hospitals, the overall temporal trends were remarkably congruent between cohorts. This suggested the presence of shared pathophysiological pathways in the terminal phase of life, despite substantial differences in patient demographics, healthcare systems, and data collection protocols (**Fig. 2**).

**Figure 2:**
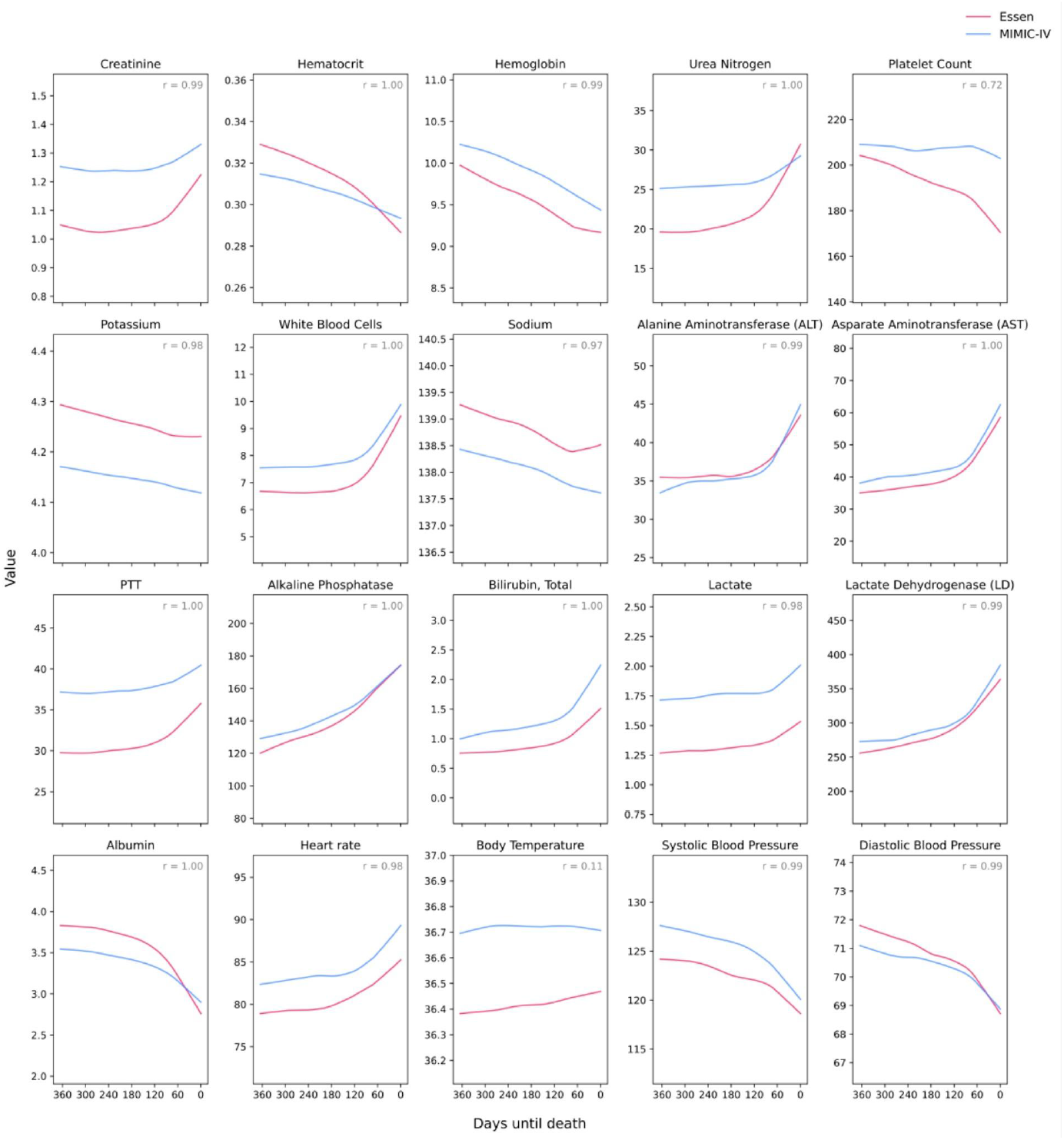
Temporal evolution of laboratory and vital sign parameters during the final year of life across the Essen and MIMIC cohorts. Trajectories of 20 clinical parameters over the last 365 days before death in hospitalized patients. Red lines represent data from the Essen cohort (Germany, *n* = 42,508). Blue lines represent the MIMIC-IV cohort (USA, *n* = 37,525). The x-axis indicates time until death (0 = day of death), while the y-axis shows the average value of each parameter over time. For each variable, daily mean values were calculated and modeled using regression-based curve fitting. Correlations were calculated using Pearson’s correlation coefficient.

### End-of-life trajectories are preserved across age, sex, and diagnoses

As our comprehensive datasets included deceased patients from two hospitals, the study population reflects the diversity of real-world clinical populations in both demographics and underlying diagnostic profiles. We therefore investigated whether the temporal trajectories observed at the population level were preserved across key clinical and demographic subgroups (**Fig. 3**).

**Figure 3:**
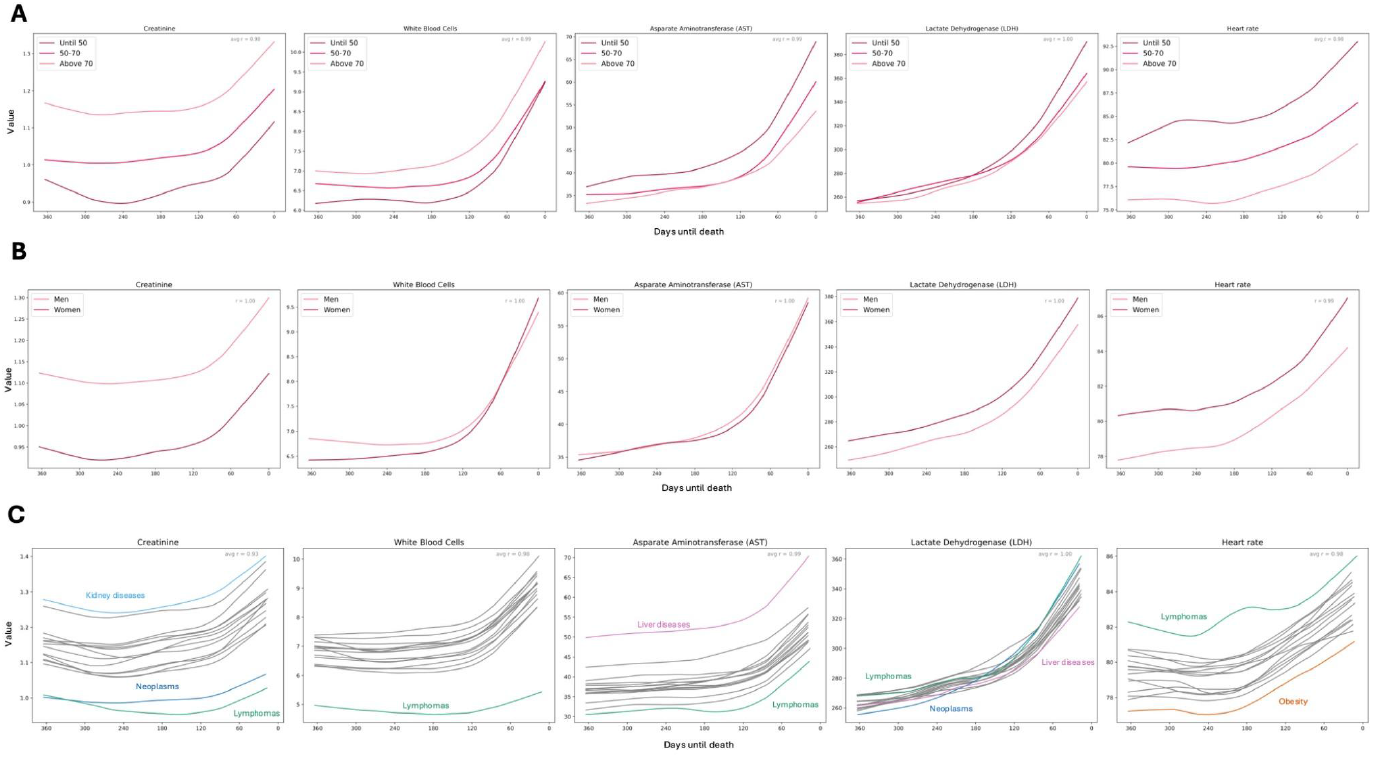
Trajectories of selected laboratory parameters by age, sex, and disease category in the internal Essen cohort. Temporal evolution of five key laboratory and clinical parameters - creatinine, white blood cell count, aspartate aminotransferase (AST), lactate dehydrogenase (LDH), and heart rate in the final year before death. The trajectories were stratified by age categories (**A**), sex (**B**), and ICD-10 diagnostic groups (**C**). The x-axis indicates time until death (0 = day of death), while the y-axis shows the average value of each parameter over time. For each variable, daily mean values were calculated and modeled using regression-based curve fitting. Correlations were calculated using Pearson’s correlation coefficient.

While absolute values of several parameters in the Essen cohort, such as creatinine, leukocytes, aspartate aminotransferase (AST), and lactate dehydrogenase (LDH), varied across subgroups, the overall direction and progression of change over time remained largely consistent. For example, as expected, older patients exhibited higher baseline creatinine levels, consistent with age-related declines in renal function. While the absolute difference in creatinine levels between older and younger patients remained stable over time, both age groups displayed similar trajectories, with pronounced elevation in the final months of life (**Fig. 3A**). Similar patterns were observed between male and female patients, who differed in baseline levels but followed parallel overall trajectories (**Fig. 3B**). Importantly, the temporal trajectories of clinical parameters were largely preserved across disease-specific subgroups (**Fig. 3C**). While we observed clinically plausible differences in absolute values, such as elevated levels of aspartate aminotransferase (AST) in patients with liver-related diagnoses, the overall end-of-life patterns remained consistent across diagnostic categories.

These findings were independently validated in the external MIMIC cohort (**Suppl. Fig. 2**).

### Multisystem dysfunction characterizes the terminal phase

To quantify the extent and overlap of dysfunction across physiological systems, we analyzed the co-occurrence of laboratory abnormalities in a subgroup of patients from our internal (n=4,579) and external (n=8,301) cohorts. Laboratory parameters were categorized into seven clinically relevant functional systems: tissue damage, coagulation, inflammation, renal function, electrolytes, hematology, and liver function. Notably, 98.3% of patients showed concurrent abnormalities in at least two of the seven functional systems, and 75.5% had concurrence in five or more systems. This underscores the predominance of multisystem dysfunction during the terminal phase (**Fig. 4A+B**). This extensive dysregulation suggests that system failure near the end of life rarely occurs in isolation, but rather as part of a broader, coordinated pathophysiological collapse.

**Figure 4:**
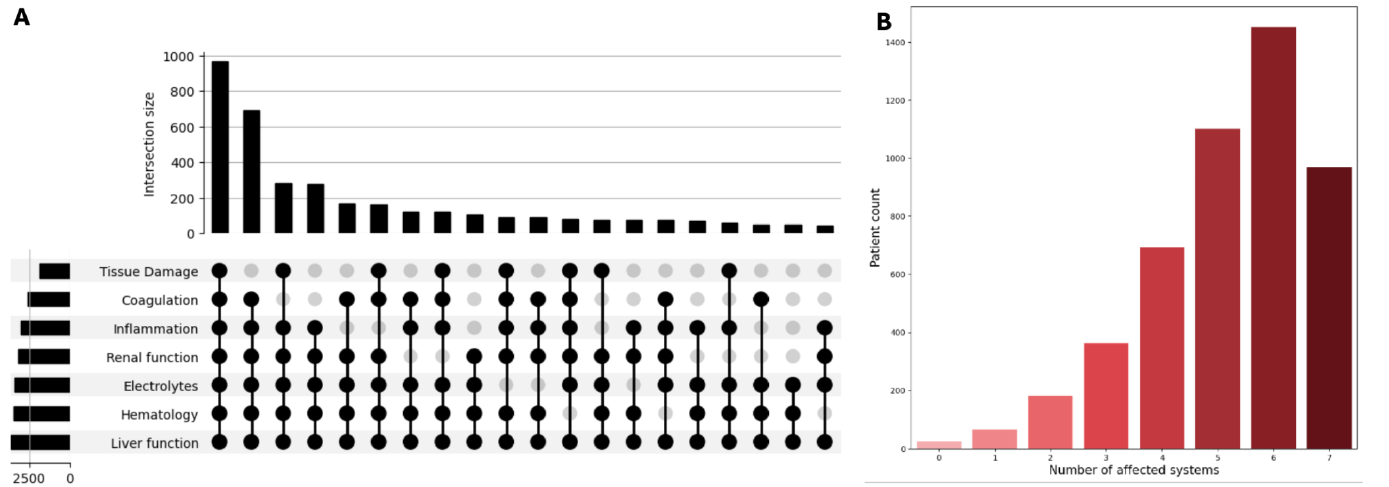
Co-occurrence and extent of laboratory abnormalities across physiological systems in the Essen cohort. **A:** UpSet plot showing the co-occurrence of significant temporal changes in laboratory parameters across seven key physiological systems: tissue damage, coagulation, inflammation, renal function, electrolytes, hematology, and liver function. Each bar indicates the number of patients with simultaneous alterations in the specific combination of systems marked by connected black dots. **B:** Distribution of the number of physiological systems affected per patient.

These findings were validated in the external MIMIC-IV cohort, where 97.8% of patients had at least two systems affected, and 54.1% had five or more systems compromised (**Suppl. Fig. 3**).

### A consistent sequence of multisystem decline precedes death

To investigate whether pathophysiological decline towards the end of life follows a structured temporal pattern, we analyzed the sequence of deterioration of physiological systems. We used the Pruned Exact Linear Time (PELT) algorithm to identify the timing of the most relevant changes in laboratory parameters across the internal and external patient cohorts.

By comparing the temporal sequence of change points in both datasets, we identified a structured pattern of system-specific changes occurring between approximately 150 and 30 days before death (**Fig. 5**). Despite some variability in individual markers, the temporal occurrence of change points was overall consistent between the internal and external cohorts (Spearman’s ρ = 0.598, p = 0.001).

**Figure 5:**
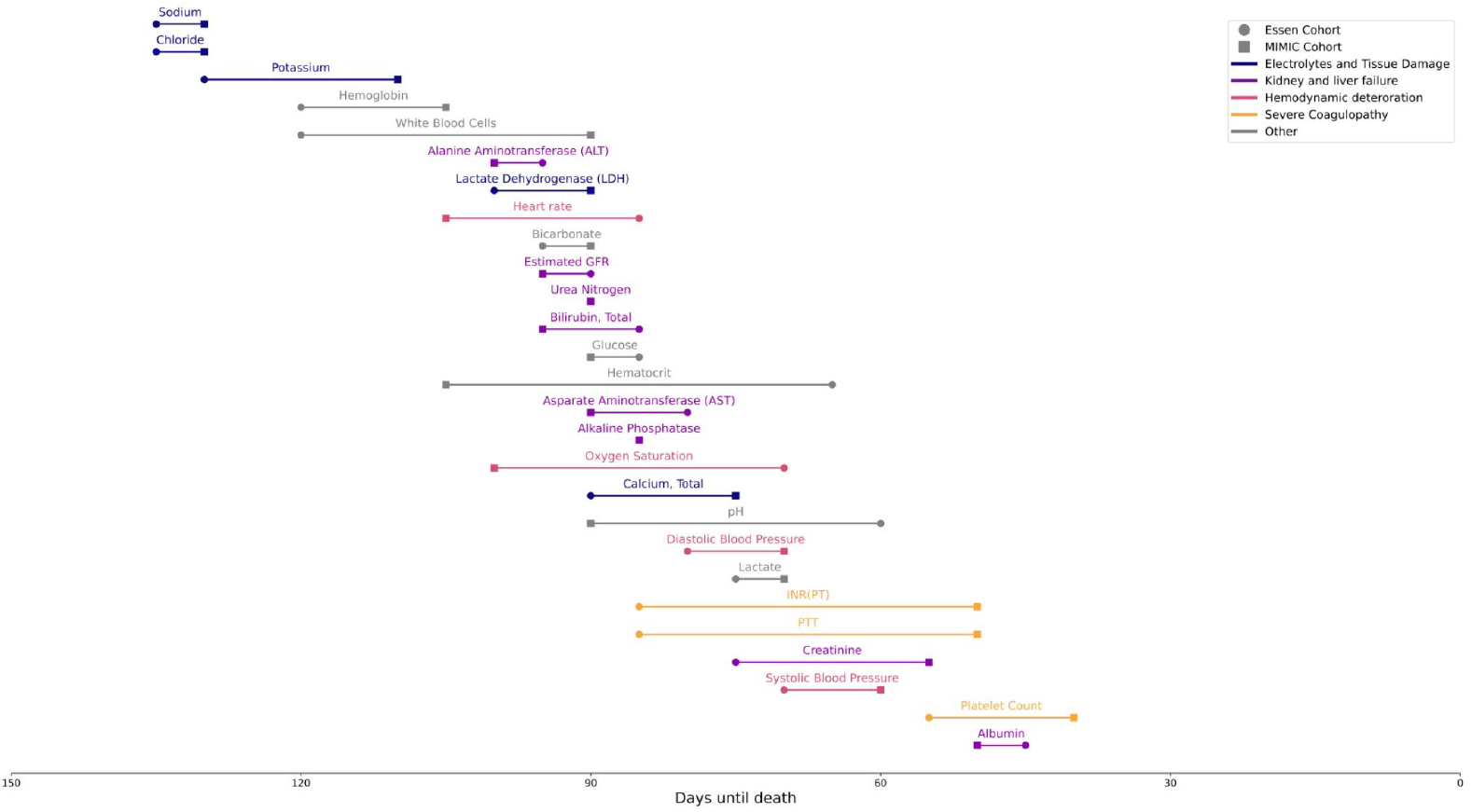
Cascade of critical alterations in diagnostic parameters preceding death. Temporal sequence of change points in 27 laboratory and vital sign parameters across the internal Essen (Circle) and external MIMIC (Square) cohorts. Change points were identified using the Pruned Exact Linear Time (PELT) algorithm. Variables are ordered by the mean timing of change across both cohorts.

The earliest changes observed were electrolyte imbalances, most notably of sodium and chloride (132.5 days before death) and potassium (120 days), signaling early systemic stress. These were followed by shifts in organ-specific markers, including liver-related (ALT), bilirubin, AST, alkaline phosphatase - 100, 90, 87.5, 85 days) and renal parameters (glomerular filtration rate [GFR], urea nitrogen, creatinine - 92.5, 90, 65 days), reflecting progressive hepatic and renal deterioration. This phase was accompanied by changes in vital signs, including heart rate (95 days) and blood pressure (systolic: 65 days, diastolic: 72.5 days), indicating a comprehensive decline in physiological stability and systemic compensation. In the terminal phase, coagulation parameters, such as INR (67.5 days), partial thromboplastin time (PTT, 67.5 days), and platelet counts (47.5 days), showed marked deviations, indicating advanced systemic failure and impending death.

### Development of a machine learning model for prediction of 90-day mortality

Building on our identification of distinct and generalizable patterns preceding death, we developed a machine learning model for the prediction of 90-day mortality. The model was trained on 80,806 patients (42,279 deceased, 38,527 non-deceased) from the internal Essen database, incorporating 27 clinical parameters per patient, including both absolute values and temporal trends derived from historical changes in laboratory results. This allowed the model to capture dynamic, time-dependent pathophysiological changes. This diagnosis-agnostic model achieved strong prediction performance when tested on the internal Essen cohort (area under the curve, AUC = 0.86; **Fig. 6A**). External validation on the MIMIC-IV cohort (211,527 patients overall, 37,969 deceased, 173,558 non-deceased) demonstrated robust model performance (AUC = 0.79; **Fig. 6B**). Global feature importance, based on mean absolute SHapley Additive exPlanations (SHAP) values, highlighted laboratory parameters and their temporal dynamics as key contributors to model predictions in both datasets (**Fig. 6C**). Notably, PTT, hemoglobin, and white blood cell counts, as well as their temporal changes (e.g., PTT_delta, hemoglobin_delta), were among the most influential features.

**Figure 6:**
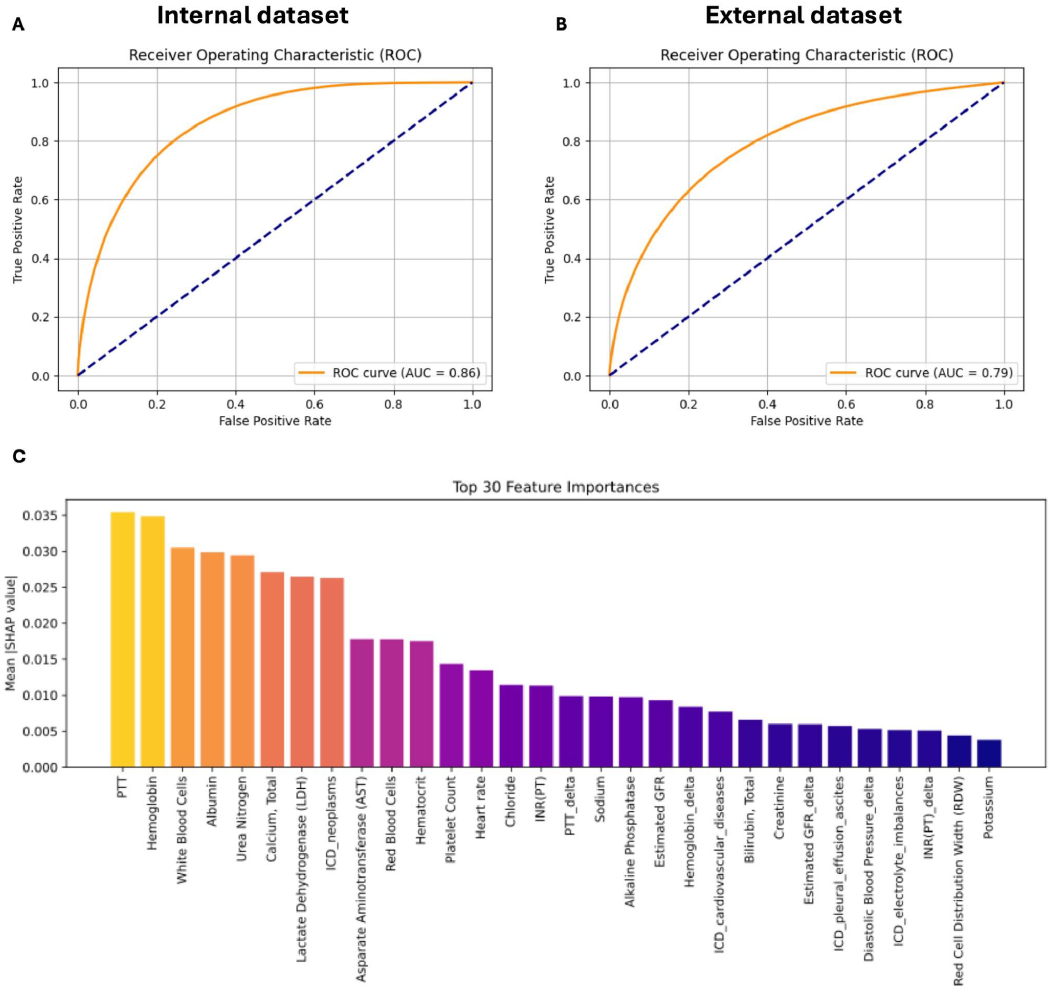
Performance and interpretability of a 90-day mortality prediction model based on clinical routine data. **A**: Receiver Operating Characteristic (ROC) curve showing the model performance when trained and validated on the internal Essen cohort (80/20 split). **B**: ROC curve of external validation using the MIMIC-IV cohort after training on the Essen cohort. **C**: Mean absolute SHAP values visualizing the global feature importance as derived from a Random Forest classifier using the SHAP TreeExplainer. The binary classifier was trained to predict 90-day mortality (0: survival, 1: death) using ICD-10 codes, laboratory values, clinical parameters, and their temporal changes (delta values) as input features.

## Discussion

Although death is a fundamental part of clinical care, its pathophysiological processes remain incompletely understood. Accurately characterizing and tracking detectable patterns that precede potentially irreversible decline is crucial for informed decision-making and personalizing care at the end of life. In this study, we systematically analyzed the trajectories of 42,508 patients in their final year of life using time-resolved, multimodal diagnostic data, and validated our findings in an external cohort of 37,525 patients. We found that physiological decline follows characteristic and systematic patterns across various primary diagnoses. This includes laboratory parameters, comorbidities, and vital signs. These conserved biomarker trajectories capture progressive multisystem deterioration and can be leveraged by machine learning models to predict end-of-life decline.

Due to data limitations, previous approaches have largely focused on short-term outcomes, specific disease groups, or limited data modalities, often lacking systematic analysis, validation, and scalability.^12–19^ In our study, we used a diagnosis-agnostic, longitudinal approach to capture generalizable, rather than disease context-specific relationships between multimodal clinical data and death. This enabled us to reveal consistent patterns of terminal decline, which are conserved across age, sex, and clinical diagnoses. Notably, the observed longitudinal laboratory trajectories were consistent with findings from other large-scale population-based studies, further reinforcing the robustness and generalizability of these trends across different clinical settings.^19^

Specifically, we found that most routine clinical markers follow distinct, startpoint- and slope-specific trajectories. While some markers, such as altered sodium and chloride concentrations, are early signs of decline, other markers, including abnormalities of PTT and albumin, are detected shortly before death. In most patients, physiological systems, such as liver and kidney, were concurrently affected, supporting the hypothesis that terminal physiological decline reflects a globally dysregulated state rather than isolated organ failure.^20,21^ However, multi-system dysfunction does not start simultaneously, but follows a cascade that reflects a stepwise, structured progression of physiological decline, in which distinct organ systems fail sequentially. Starting with signs of electrolyte imbalances and general tissue damage, the deterioration progresses to hepatic and renal dysfunction, ultimately culminating in coagulation failure. Consistent with earlier findings, our results confirm that end-of-life involves failure across multiple organ systems.^20,21^ The identification of distinct and conserved patterns of terminal decline enabled us to develop a prediction model for estimating 90-day mortality risk. Training the model on large-scale internal data, we achieved strong predictive performance on both the internal (AUC: 0.86) and external validation datasets (AUC: 0.79). These results confirmed that the model generalizes well across geographically, structurally, and clinically diverse patient populations. It highlights its potential applicability in a wide range of clinical settings and healthcare systems.

Understanding the model’s decision-making process is essential for the responsible and widespread application of machine learning in clinical practice^22,23^. To this end, we used explainable AI (xAI) techniques to assess the contribution of each individual parameter. ^24,25^ Notably, parameters with pronounced changes during the final year of life, such as PTT, hemoglobin, and white blood cell counts, played a central role in the model’s decision-making. Additionally, 5 of the 30 most impactful parameters were ‘delta parameters’ reflecting time-dependent changes. This underscores the important prognostic value of dynamic trajectories in addition to single-time-point measurements.

Nevertheless, our study is not without limitations. First, although we have analyzed large, real-world datasets from two independent hospital systems, both cohorts are limited to hospitalized patients, which may not fully capture trajectories in patients who die outside the hospital or receive care exclusively in a community setting. Furthermore, despite the generalizability of our findings across different institutions, retrospective analyses are inherently limited by potential biases in data availability, coding practices, and unmeasured confounders.^26,27^ Also, while we incorporated temporal dynamics through ‘delta parameters’, the resolution of historical data varies across patients, potentially impacting the model’s ability to capture early changes. Future work should explore the integration of patient-reported outcomes to further support patient-centered treatment decisions and individualized care planning.

In summary, our study demonstrates that widely available makers from routine clinical care contain highly conserved and generalizable signals of terminal pathophysiology. By developing a prediction model for 90-day mortality, our study contributes to a data-driven approach aiming to support informed decision-making between patients and clinicians, shifting the perspective from reactive care to anticipatory and proactive care planning.

## Methods

### Study design and data processing

We utilized two patient cohorts: one from University Hospital Essen, Germany, and another from the publicly available MIMIC-IV database.^9–11^ The data from Essen were retrieved via the Smart Hospital Information Platform (SHIP) of University Hospital Essen, which integrates structured clinical data from various hospital subsystems using the FHIR standard. The study was approved by the Ethics Committee of the Medical Faculty of the University Duisburg-Essen (No. 22-10881-BO). The requirement for written informed consent was waived due to the retrospective design of the study and the deidentification of data.

To enable cross-cohort comparability, we identified the most frequently measured laboratory parameters in both datasets and harmonized unit discrepancies between them. For descriptive analysis, we included only laboratory measurements recorded within 365 days prior to death. Patients younger than 18 years and those with fewer than five laboratory measurements across the entire follow-up period were excluded due to limited interpretability. Extreme outliers were removed by excluding values below the 1st and above the 99th percentile, followed by z-score filtering with a threshold of ±3 (**Suppl. Figure 1**).

### Diagnosis Grouping

ICD codes were truncated to the first three characters and subsequently grouped into clinically meaningful diagnostic categories:

- C00-C81: Neoplasms
- I10–I15: Hypertensive Disorders
- I20–I52: Cardiovascular Diseases
- E10–E14: Diabetes Mellitus
- E66, E78: Obesity
- E86–E87: Electrolyte Imbalances
- J09–J18: Pneumonia
- J40–J47: Chronic Respiratory Diseases
- J90–J94 + R18: Pleural Effusion and Ascites
- K70–K77: Liver Diseases
- N00–N19: Renal Diseases
- A40–A41 + R65: Sepsis
- C81–C96: Lymphomas
- D50–D64: Hematologic Disorders
- R57: Shock

ICD-9 codes present in the MIMIC dataset were mapped to ICD-10 codes using a crosswalk and categorized identically.

### Descriptive Analysis

Laboratory time series were merged with ICD code groupings, allowing patients to appear in multiple diagnostic categories. Additional variables such as age and sex were integrated for subgroup analyses. Laboratory trends over time were visualized across ICD groups, age, and sex strata using regression-based curve fitting.

To detect significant monotonic changes in laboratory values, we applied the Mann-Kendall trend test (pymannkendall) to each patient’s individual time series. Prior to analysis, comprehensive preprocessing was performed, including outlier removal and the aggregation of ICD codes into medically meaningful diagnostic groups. To ensure temporal resolution and comparability, only patients with at least four measurements in each of the investigated laboratory categories were included, yielding a final cohort of 4,579 patients. Statistically significant trends were identified and categorized into seven functional groups:

- Coagulation: INR, PTT
- Liver function: ALT (GPT), AST (GOT), Total Bilirubin, Total Protein
- Electrolytes: Potassium, Sodium, Total Calcium, Chloride
- Renal function: Creatinine, Urea Nitrogen, Estimated GFR
- Hematology: Hematocrit, Hemoglobin, Erythrocytes, Platelet Count
- Inflammation: CRP, Leukocytes
- Tissue Damage: LDH

The frequency and overlap of significant trend patterns were visualized using an UpSet plot (Figure 4, UpSetPlot library).^28^ We validated these results on our MIMIC-IV cohort, applying the same processing steps (N=8,301). Due to insufficient measurements, we had to exclude estimated GFR, total protein, and CRP, resulting in 17 features for the analysis.

To identify non-monotonic patterns, we applied the PELT algorithm from the ruptures Python library^29^ to detect change points in the smoothed laboratory time series after outlier removal. The first detected change point for each parameter was used to construct a temporal cascade of changes, enabling comparison between the Essen and MIMIC cohorts (Figure 5). The relative ordering of change points across cohorts was then compared using Spearman correlation.

### Predictive Modeling

In addition to the patients included in the descriptive analysis, those who died within 365 days of their last recorded measurement, we incorporated a control group of patients who did not die within this time frame for the development of the predictive model. This group was randomly sampled from the same underlying datasets, ensuring comparable data quality and measurement density. By including both terminal and non-terminal cases, we aimed to increase the robustness and generalizability of the model. This design enabled the model to distinguish between typical fluctuations in clinical parameters and those indicative of impending physiological collapse, thereby enhancing stability and reducing the risk of overfitting to end-of-life–specific trends.

For predictive modeling, the data were transformed into a wide format, where each row represented a patient-day with available laboratory values. Missing values were backfilled per patient by looking up to 10 days into the past. In addition to raw laboratory values, we calculated a delta feature for each lab parameter, representing the change from the previous measurement. This delta was normalized by the number of days since the last measurement to account for varying measurement intervals. Missing delta values were kept as NaN to allow for consistent downstream imputation.

Only features with less than 70% missing values in the Essen cohort were retained. For remaining missing values, k-nearest neighbor imputation (scikit-learn package^30^) was applied if no prior measurement was available. ICD codes were accumulated over time to reflect chronic conditions, one-hot encoded, and merged with laboratory values by patient and timestamp

The dataset was split into training and test sets based on patients (80:20 split). The binary outcome was defined as death within the next 90 days (class 1) vs. survival (class 0). All features were standardized using the StandardScaler from scikit-learn. A Random Forest from the publicly available scikit-learn library classifier was trained on the processed data to predict 90-day mortality. The shown ROC curves were also calculated using the scikit-learn library.

To interpret the model and understand the contribution of individual features to the prediction, we calculated SHAP (SHapley Additive exPlanations)^31^ values for 200.000 test samples using the ForestExplainer module tailored for tree-based models. This allowed us to derive a global measure of feature importance by quantifying how much each variable contributed, on average, to the model’s output. The SHAP framework offers consistent and locally accurate attributions, making it particularly suitable for interpreting complex ensemble models such as Random Forests.

### External Validation

When validating the model results on our MIMIC cohort, we performed the same preprocessing steps as with the Essen cohort. While the imputer was fitted on the complete dataset in Essen, we applied it to a randomly selected subset of 10,000 patients from the MIMIC cohort.

### Statistics

Statistical analyses and model development were conducted using Python (v3.10) and associated open-source packages. Data preprocessing and statistical testing were conducted without randomization or blinding of the investigators to cohort membership.

Descriptive statistics and trend analyses were performed separately for each cohort. Outliers were removed by excluding values below the 1st and above the 99th percentiles, followed by z-score filtering (threshold ±3). Smoothening of the trajectories was performed using the statsmodels Python package^32^. The correlations shown in Figure 1 and 2 were calculated using Pearson’s correlation coefficient, implemented via the scipy Python package^33^. The Mann-Kendall trend test from the pymannkendall Python package^34^ was applied to patient-level time series to detect significant monotonic trends in laboratory values over time. Statistical significance was defined as *p* < 0.05. The analysis was limited to patients with ≥4 timepoints per laboratory parameter. For non-monotonic temporal changes, change point detection was performed using the PELT algorithm from the ruptures Python package, applied to smoothed lab trajectories. The temporal ordering of change points was compared between cohorts using Spearman rank correlation from the scipy Python package^33^.

For predictive modeling, the outcome was defined as mortality within 90 days of a given measurement day. Data were transformed into patient-day format, and missing values were imputed using k-nearest neighbor imputation (scikit-learn^30^). Delta features were computed as change per day since the previous measurement. Feature selection was based on missingness thresholds and domain relevance. Random Forest models were trained using the scikit-learn implementation with default hyperparameters unless otherwise specified. Model performance was assessed on a held-out test set using ROC curves and AUC metrics.

Model interpretability was evaluated using SHapley Additive exPlanations (SHAP) values for 200,000 randomly sampled patient-days from the Essen cohort. SHAP values were computed using the TreeExplainer from the shap Python package^35^ to assess the global and local impact of individual features on model predictions.

## Data Availability

Data from University Hospital Essen cannot be shared with investigators outside the institution without consent.

https://physionet.org/content/mimiciv/3.1/

## Acknowledgments

The data for this project was provided by the Smart Hospital Information Platform (SHIP), managed by the Data Integration Center at the University Medicine Essen. SHIP serves as a comprehensive digital health platform for integrating data from all major clinical subsystems using a holistic FHIR-based approach. It enables the purification, analysis, distribution, and visualization of clinical data.

## Funding

J.Keyl is supported by a German Research Foundation (DFG)-funded clinician scientist program (FU 356/12-2).

### Author contributions

Conceptualization: JKeyl, PK, JKleesiek

Methodology: JKeyl, PK, JKleesiek

Formal analysis: JB, JKeyl, PK, TL

Investigation: all authors

Data acquisition and evaluation: JB, JKeyl, TL, DF-S, MW, FK, MS, SH, PK, JKleesiek

Data curation: JB, JKeyl, TL

Writing - original draft: JB, JKeyl, PK, JKleesiek

Writing - review and editing: all authors

Supervision: JKleesiek, PK

