## Supplementary Material for "Characterizing and Predicting End-of-Life Patient Trajectories Using Routine Clinical Data"

### Supplements

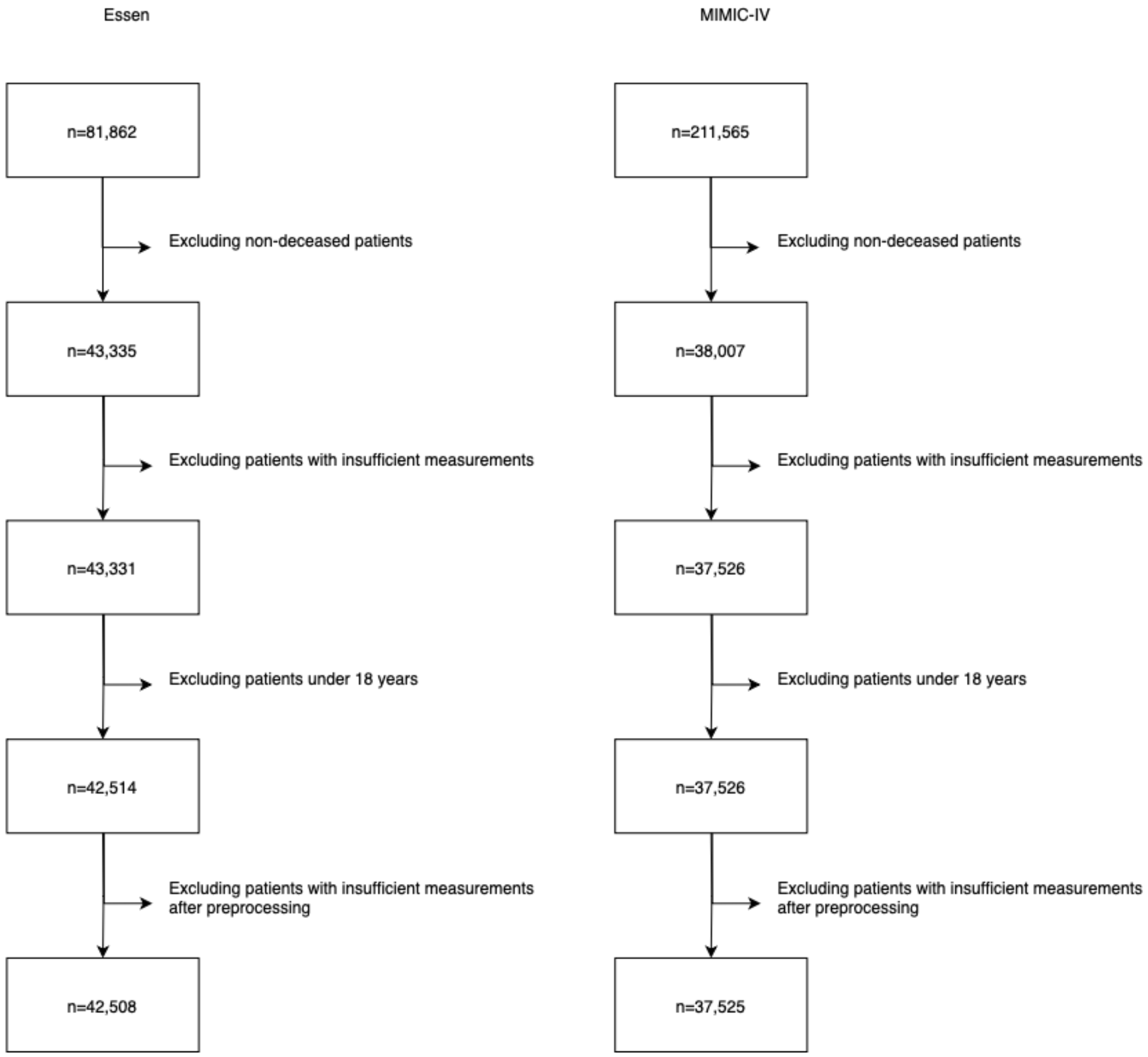


**Supplement Figure 1: Preprocessing steps**
Preprocessing for both the internal Essen cohort, as well as for the external MIMIC cohort for the descriptive analysis. Patients with insufficient measurements, of age under 18 years and outliers were removed.

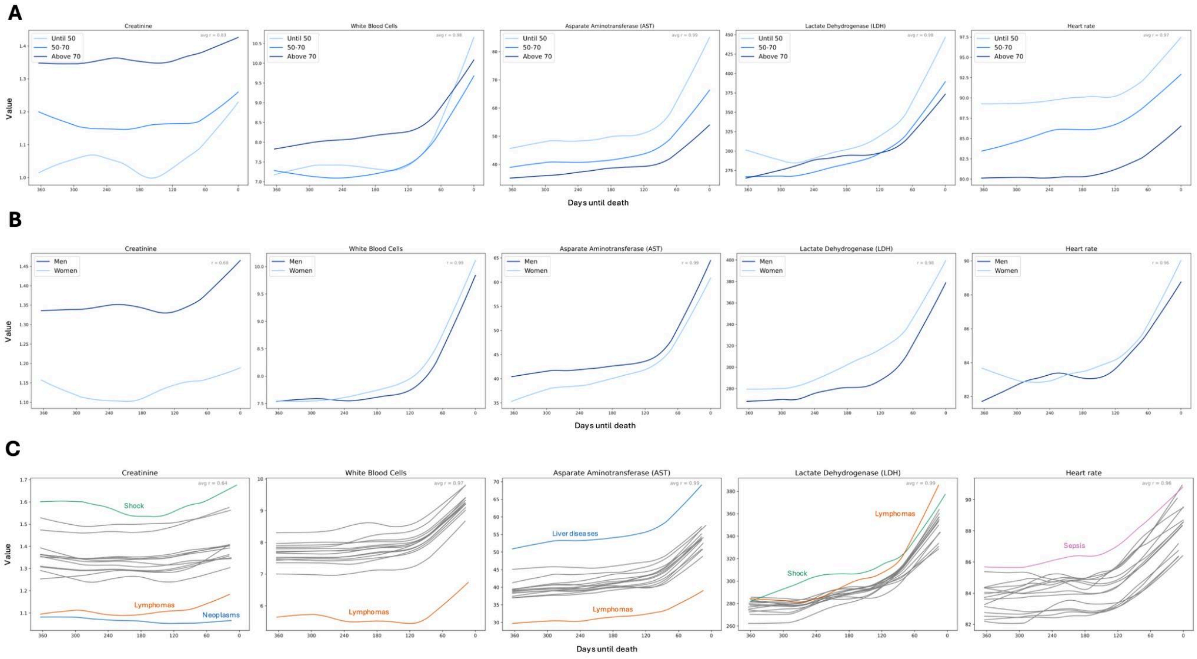


**Supplement Figure 2: Trajectories of selected laboratory markers by age, sex, and disease category in the external MIMIC cohort.**

Temporal evolution of five key laboratory and clinical parameters - creatinine, white blood cell count, aspartate aminotransferase (AST), lactate dehydrogenase (LDH), and heart rate in the final year before death. The trajectories are stratified by age categories (**A**), sex (**B**) and ICD-10 diagnostic groups (**C**). The x-axis indicates time until death (0 = day of death), while the y-axis shows the average value of each parameter over time. For each variable, daily mean values were calculated and modeled using regression-based curve fitting. Correlations were calculated using Pearson's correlation coefficient.

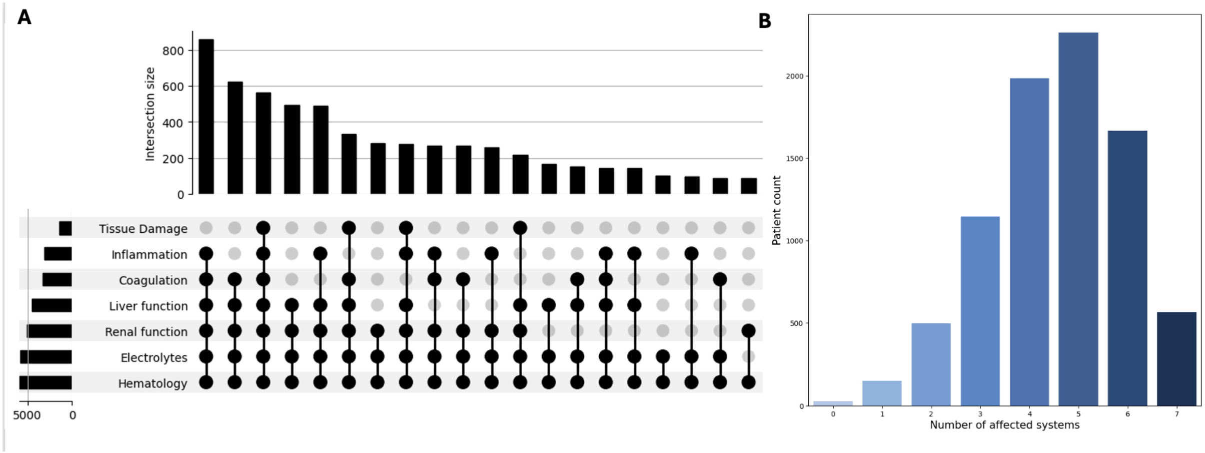


**Supplement Figure 3: Co-occurrence and extent of laboratory abnormalities across physiological systems in the external MIMIC cohort.**

**A:** UpSet plot showing the co-occurrence of significant temporal changes in laboratory markers across seven key physiological systems: tissue damage, coagulation, inflammation, renal function, electrolytes, hematology, and liver function. Each bar indicates the number of patients with simultaneous alterations in the specific combination of systems marked by connected black dots. **B:** Distribution of the number of affected systems per patient.
